# Refining the Analysis of Multidimensional Psychometric Data: A Geometric Distance Approach

**DOI:** 10.1101/2021.10.14.21265002

**Authors:** Dan W. Joyce, Nicholas Meyer

**Affiliations:** Department of Psychiatry, University of Oxford and NIHR Oxford Health BRC, Warneford Hospital, Oxford, OX3 7JX; Institute of Psychiatry, Psychology and Neuroscience, King’s College London, 16 De Crespigny Park, London, SE5 8AF

## Abstract

Validated instruments such as questionnaires, patient-reported outcome measures and clinician-rated psychopathology scales, are indispensable for measuring symptom burden and mental state, and for defining outcomes in both psychiatric practice and clinical trials. Most often, the values on the instrument’s multiple items (dimensions) are added to derive a single, univariate (scalar) sum-score. Although this approach simplifies interpretation, there are always many possible combinations of individual items that can yield the same sum-score. Two patients can therefore obtain identical scores on a given instrument, despite having very different combinations of underlying item scores corresponding to different patterns of clinical symptoms. The same is also true when a single patient is measured at two different time points, where the resulting sum-scores can obscure changes that may be clinically meaningful.

We present an alternative analytic framework, which leverages geometric concepts to represent measurements as points in a vector space. Using this framework, we show why sum-scores obscure information present in measurements of clinical state, and also provide a straightforward algorithm to mitigate against this problem. Clinically-relevant outcomes, such as remission or patient-centered treatment goals, can be represented intuitively, as reference points or ‘anchors’ within this space. Using real-world data, we then demonstrate how measuring the relative distance between points and anchors preserves more information, allowing outcomes such as proximity to remission, to be defined and measured.

## 1 Introduction

A patient’s **clinical state** is the constellation of signs and symptoms for a disorder at a certain point in time. In clinical practice and trials, clinical states can be measured using psychometric instruments such as clinician or patient-reported scales - for example, the Positive and Negative Symptom Scale (PANSS) (Kay et al., 1987) for psychosis, Beck Depression Inventory (BDI-II) (Beck et al., 1996a) for depression and the Generalised Anxiety and Depression (GAD7) (Spitzer et al., 2006) scale for generalised anxiety disorder. Some common questions we ask of such instruments are:

1. how *severe* is the disorder at the time of measurement?
2. can we observe a *change* between serial measurements as the result of an intervention?
3. does the instrument identify *caseness* for a patient and a disorder?

For the first use case, most commonly, we use the sum of the instruments’ individual items to arrive at a continuous, scalar, univariate measure of severity. For brevity, we will refer to this as the **total sum score** (TSS) for a scale or instrument. For example, a patient with a TSS of 25 on the BDI-II has depression of “moderate” severity (Beck et al., 1996b) and we conclude that another patient with a score of 15 has a *lower* symptom burden. To answer the second question, the simplest measure of change is the difference in TSS between a baseline and later time point. Alternatively, we could use the percentage change in TSS from baseline (Leucht et al., 2009) or in the case of multi-arm controlled trial outcomes, use a regression model of the later TSS with the baseline score as a covariate (Vickers, 2001). For the third question, when a suitable instrument is used in a diagnostic process, we may use the TSS and an operating threshold/decision rule e.g. a GAD7 score exceeding 10 suggests a diagnosis of generalised anxiety disorder (Spitzer et al., 2006) or we might select subsets of items from the scale e.g. identifying patients with predominantly positive symptoms of psychosis, or defining criteria for remission from psychosis using subsets of items in the PANSS scale (Andreasen et al., 2005). Also, we can look for patterns or clusters within a sample using dimensionality reduction methods such as latent structure analysis, the PANSS ‘five factor’ model being an example (White et al., 1997). We might also use subsets or individual items from a scale as covariates or independent variables in analyses of other dependent variables of interest.

In psychiatric clinical trials, scales are often used as primary endpoints; in such cases, the TSS is a **composite** variable where “The method of combining the multiple measurements should be specified in the protocol, and an interpretation of the resulting scale should be provided in terms of the size of a clinically relevant benefit” (ICH Expert Working Group, 1998). TSSs provide convenience and tractability in that they a) avoid specifying multiple outcomes which obviates the need for Type I error adjustment (ICH Expert Working Group, 1998), b) are often convenient for clinical understanding (because it is easy to relate to e.g. a BDI-II score of 25 indicating a moderate degree of symptoms of depression in contrast to a score of 10 indicating minimal burden) and c) can be easily dichotomised to define response to treatment, for example defining a 50% reduction in the overall symptom burden – although these approaches have been criticized for consequent loss of statistical information (Senn and Julious, 2009, Vickers et al. (2008)).

Although these methods are convenient, by *only* employing a composite variable derived by computing the arithmetic sum (the TSS) over the whole instrument, we also risk overlooking meaningful structure present in data from multivariate scales.

In the applied statistics and trial design literature, continuous composite scores (like the TSS) have been discussed in terms of statistical efficiency; for example, for two variables *X* and *Y* (e.g. items from a scale), the effect size of the sum (*X* + *Y*) can be *greater* or *less* than the individual effect sizes of X or Y depending on their correlations and the ratio of the individual effect sizes (Liu-Seifert et al., 2017). These results translate into trial designs: in cases where a treatment only affects component *X or Y*, using the sum (*X* + *Y*) would require 2–4 times the sample size to maintain the same statistical power (Troy and Simmons, 2020). To quote (Prieto-Merino et al., 2013): “The main objection to the use of composite outcomes is that, if the treatment has different effects on the different components of the outcome, the net effect on the composite outcome is difficult to interpret”.

In the psychometrics literature, there has been a focus on the combinatorics of syndrome definitions – the multiple arrangements or combinations of items to meet a certain criteria for diagnosis. For example, there are 277 unique combinations of DSM-5 criteria for major depressive disorder and 341,737 different combinations for the melancholic specifier (Fried et al., 2020). Further, composite measures have been argued to “carry information about the general psychopathological load of a particular person, but… the approximation may be fairly rough and that summing symptoms may ignore important information” (Fried and Nesse, 2015b). In 3703 patients from the STAR*D trial, 1030 unique profiles of symptom combinations were identified, of which 864 were present in only 5 or fewer patients (Fried and Nesse, 2015a). In this situation, establishing whether or not a patient meets operationalised diagnostic criteria is analogous to scoring a collection of binary items in a multidimensional scale.

We pursue an alternative (but complimentary) approach, which leverages geometric concepts to partially preserve the structure inherent in data from multivariate instruments, and delivers meaningful clinical endpoints and measures of overall severity. We propose that a measurement of clinical state from a multi-item instrument can be viewed as a point or location in a multidimensional normed vector space, which introduces concepts of **distance** and **similarity** between states. We illustrate how composite scores like the TSS are a specific example of distance measurement, and develop the central ideas in a graphical and tutorial way on a simple two-item instrument. We then present theoretical results for larger scales/instruments, with a greater number of items, and present an algorithm for computing composite scores that allow us to build-in clinically meaningful ‘anchors’ for computing composite scores. We conclude by showing how similar results manifest in real data using the PANSS data from the CATIE trial (Lieberman et al., 2005) and show examples of how to apply anchoring for composite scores to concepts of remission and treatment refractoriness.

## 2 An Explosion of Combinations

When clinical state is measured by a multi-item instrument/scale, there are many possible combinations of those items which yield the same TSS. This means that two patients with different clinical states are “mapped” to the same TSS. Consider an instrument with only two items, such as the PHQ2 scale (Kroenke et al., 2003) for depression, which measures two items over the preceeding two weeks: i) little interest or pleasure in doing things and ii) feeling down, depressed or hopeless. We will abbreviate and label these two items Anhedonia and Mood respectively. Each item is ordinally scored 0, 1, 2 and 3 representing “not at all”, “several days”, “more than half the days” and “nearly everyday” respectively, and ordinal levels for all items (axes, dimensions) must be meaningfully ordered such that a level of 0 on an item is “strictly less than” 1, which is strictly less than 2 and so on.

In Figure 1 (left-panel) we plot the ordinal levels for Anhedonia and Mood in ascending order along orthogonal (perpendicular) axes; all combinations of the levels of Anhedonia and Mood are unique points in the plane and by counting these points we conclude there are 4^2^ = 16 possible combinations, or unique clinical state measurements. The TSS corresponding to a given unique point is shown as a colour and the co-ordinates of each location are shown as e.g. (1, 3) interpreted as a score of 1 on the Anhedonia axis and 3 on the Mood axis.

**Figure 1:**
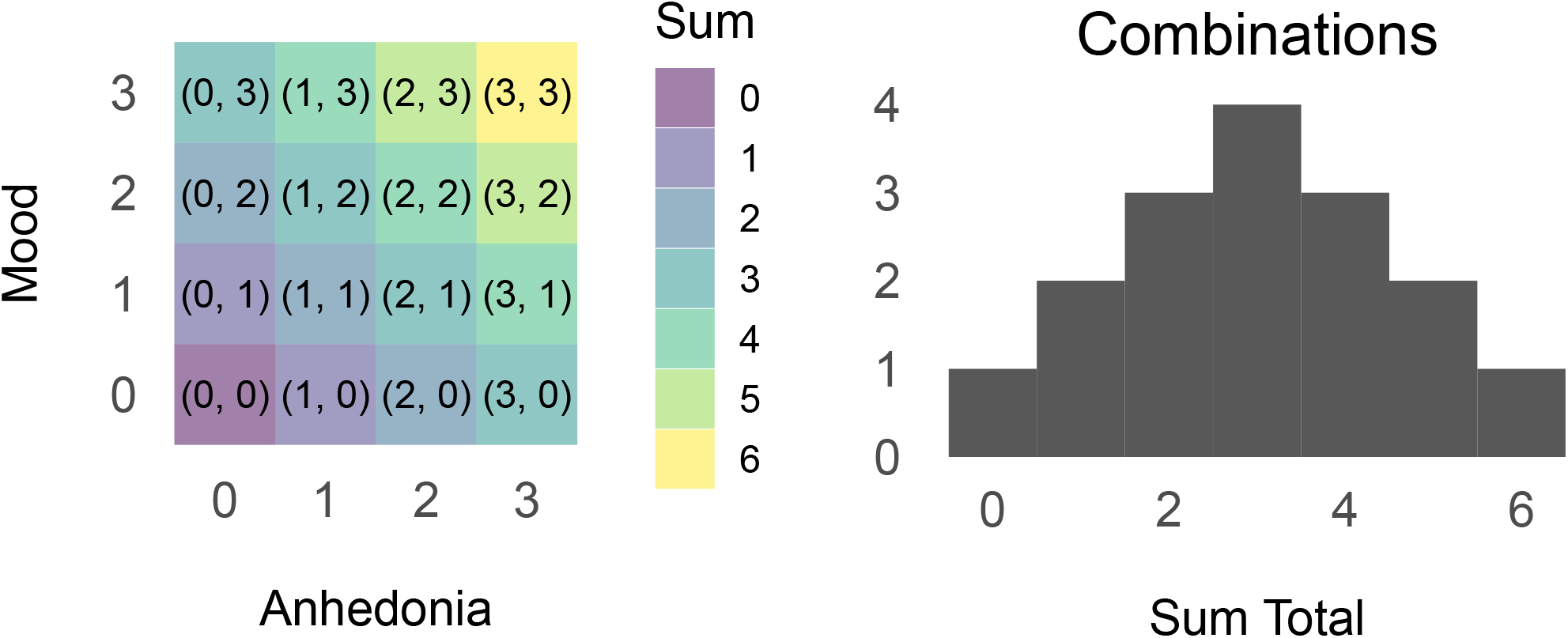
Possible Combinations and Clinical States for a Two Item Instrument

Assigning each item (Anhedonia, Mood) in the instrument to an orthogonal axis (or dimension) in this way forms a **coordinate** or **vector space** (Strang, 2006; Rudin, 1976). **We will denote the number of axes (analogously, dimensions or items in the instrument) by *d* and call this the native** space for the instrument. In this simple example, we have *d* = 2 items so we can represent states as points on a two-dimensional plane. For *d* = 3 we would need three axes (a cuboid) and visualising *d* > 3 becomes impossible without employing a dimensionality reduction or projection method (i.e. methods that essentially compress or map the native higher-dimensional space into a two-dimensional plane with the hope that spatial/geometric relationships in the native space are preserved in two-dimensions).

The notation we will use is that each location/point in the native space is a vector (an ordered list of elements or numbers) which in our two-dimensional example might be:

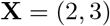

representing a score of 2 on Anhedonia, 3 on Mood and with TSS = 5. The same principle applied to the GAD7 would result in, for example:

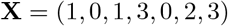

Here, the first element is 1, representing the first GAD7 item “Feeling nervous, anxious or on edge” having value 1 (“several days”). The fourth element represents the item “Trouble Relaxing” and scores 3 (“more than half the days”). The individual elements (items on the instrument) in the vector are identified by subscripted lower-case letters *x*_*i*_; for example, *x*_3_ and *x*_7_ refer to the third and seventh elements of **X** respectively. For a measurement using PANSS, we would have a vector of 30 elements for each measurement **X** = (*x*_1_, *x*_2_, …, *x*_30_).

In general, therefore, for an instrument with *d* items each assuming one of *p* ordinal levels we have a total of *p*^*d*^ possible combinations (clinical states or unique locations/points in the native space). For example, the GAD7 has *d* = 7 items scored between 0 and 3 (thus, *p* = 4 ordinal levels) giving 4^7^ = 16,384 unique combinations. The BDI-II has *d* = 21 items each with *p* = 4 ordinal levels, yielding 4^21^ ≈ 4.39 × 10^12^ combinations. The PANSS has *d* = 30 items each scored between 1 and 7 (*p* = 7) resulting in 7^30^ ≈ 2.25 × 10^25^ combinations. As a scale comparator, the time elapsed since the big bang is estimated to be of the order 10^17^ seconds and there are of order 10^10^ neurons in the human brain.

However, more important than combinatorial explosion as a function of dimensions and levels is how we count the *different combinations* that result in the *same* TSS. The TSS is simply the sum of the elements in the vector (coordinates) for a given point. For example, on the PHQ2, a point **X** = (2, 3) has a TSS of 2 + 3 = 5. In the GAD7 example above, the point **X** = (1, 0, 1, 3, 0, 2, 3) has TSS = 1 + 1 + 3 + 2 + 3 = 10.

Returning to the example in Figure 1, we have Anhedonia and Mood ranging from 0 to 3. The coloured ‘cells’ show the TSS (Anhedonia + Mood) at each of the 16 possible locations. For a TSS of 0, there is a single and unique state or point which is (0, 0). For a TSS of 1, there are *two* possible points, (1, 0) and (0, 1). For a TSS of 3, there are *four* possible, unique locations; the diagonal cells running from the top-left to the bottom-right of the figure above. In this example, we can easily select any TSS (from 0 to 6) and then count the number of different combinations of Anhedonia and Mood that gives rise to that sum. For the PHQ2 example, a histogram of the number of different combinations (unique locations/points) that give rise to every TSS is shown in the right panel of Figure 1.

For the PHQ2, manually enumerating the possible combinations of Anhedonia and Mood items which result in any sum total is straightforward. The more general version of this problem (enumerating all possible combinations of *d* items that yield a given TSS), however, is the **subset sum problem** for which there are no tractable analytical solutions (Garey and Johnson, 1979). We can use brute-force enumeration methods but only for relatively small examples. Applying this same approach to the GAD7, we have *d* = 7 items, and *p* = 4 ordinal levels for each item and enumerating all combinations by brute-force we arrive at the histogram shown in Figure 2. There are 2128 different combinations of GAD7 items (clinical states) that give the *same* TSS of 10 and similarly for a TSS of 11 (i.e. the mode of Figure 2). This brute-force method applied to only the *positive* subscale of PANSS (*d* = 7 items with *p* = 7 levels) yields a similarly-shaped histogram but where there are 60,691 unique combinations (points in the native space) which give rise to the *same* (modal) total sum score of 27.

**Figure 2:**
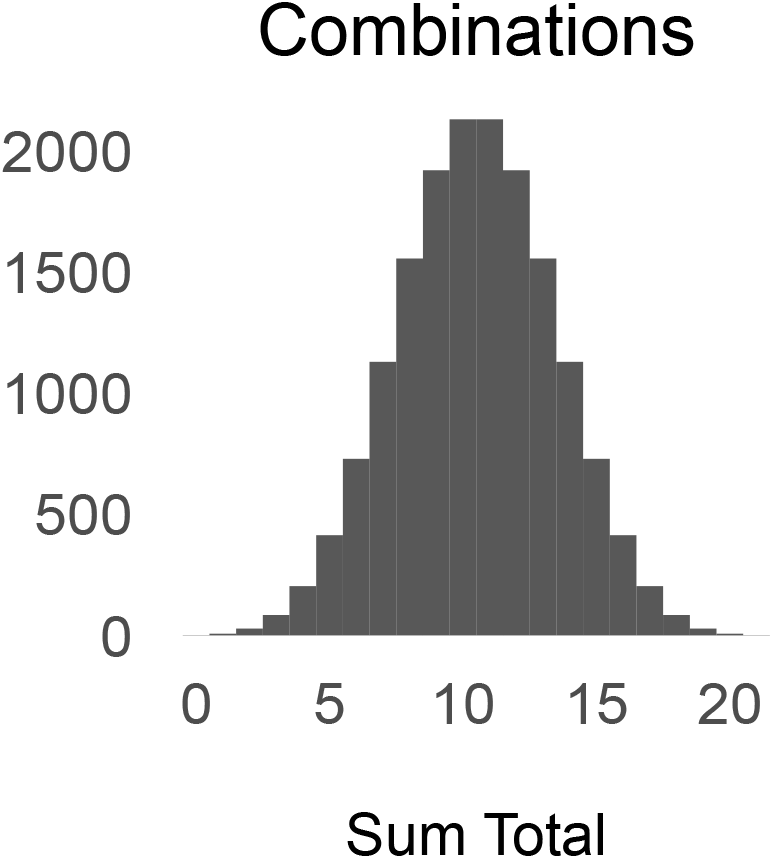
Combinations for each Sum Total on the GAD7

This demonstrates that for a **multi-dimensional** instrument (GAD7, PANSS, BDI-II etc.), summarising by computing the TSS obscures many different combinations of individual item scores and therefore, the patterns of symptoms/signs measured by the instrument. In the case of the PHQ2 example, we can see that for a TSS of 3, the points (0, 3) and (3, 0) represent completely opposing patterns of symptoms.

## 3 Measuring Similarity

In the GAD7 example, for a TSS of 10, there were 2128 different and unique locations in the native spaces. We choose two of these points and their vector representations are:

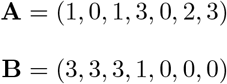

Assume that these are GAD7 measurements from two patients, A and B – they are clearly different on most items. Contrast this with the following two patients, selected from the 1128 combinations that obtain a TSS of 7:

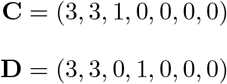

Patients C and D differ only by 1 unit on the 3rd and 4th elements. For patients A and B, the TSS fails to capture the obvious and marked differences in their respective clinical states. For patients C and D, the sum score adequately captures that they are similar.

Below, we show how introducing the notion of similarity or **distance** between vectors (points, locations or clinical states in the native space) enables us to circumvent some of the problems with TSSs and at the same time, devise a meaningful univariate or scalar summary. The approach uses standard mathematical tools that treat multi-item instruments as vector spaces and then we define **norms** and **distance** metrics (Rudin, 1976).

Visualising vectors (and spaces) in three or more dimensions is difficult, so we will use the two-dimensional PHQ2 as a concrete example throughout (i.e. we have *d* = 2 dimensions each with *p* = 4 ordinal levels), but we stress that all the principles generalise to higher-dimensional scales such as GAD7 and PANSS without modification.

In Figure 3 we identify four hypothetical patients A, B, C and D (but note, instead of being a cross-section of 4 patients, the following arguments apply equally to four serial measurements of one patient over time). Patients A and B are similar in that they have mild burden of Anhedonia symptoms and moderate-severe Mood symptoms. Patient C has *only* a severe burden of Anhedonia symptoms, but is unaffected by Mood symptoms and patient D is severely affected by both Anhedonia *and* Mood symptoms. It is clear that patient C is very different from D as well as A and B; patients A and B are quite similar, perhaps to the point where there is no meaningful clinical difference between them.

**Figure 3:**
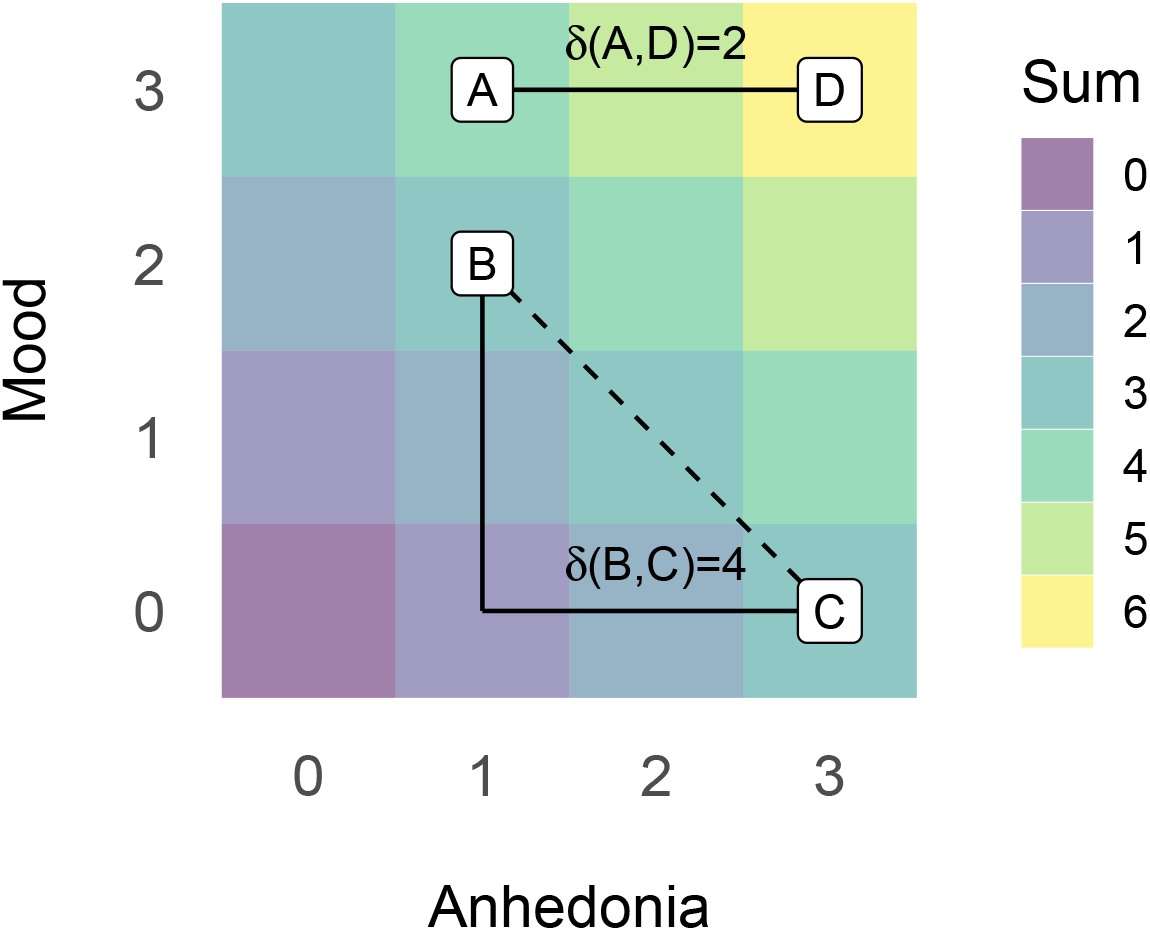
Total Sum Scores and Distances: Colours represent TSS; solid lines represent examples of rectilinear distances between patients

If we compute the TSS for each patient we find:

| Patient | TSS |
| --- | --- |
| A | 4 |
| B | 3 |
| C | 3 |
| D | 6 |

Notice that:

- Patients B and C **both** score 3, despite their obvious difference in terms of burden of Anhedonia / Mood symptoms shown geometrically in Figure 3
- Patients A and B score 4 and 3 respectively, which preserves some notion of the geometric proximity in Figure 3
- Patient D scores 6 so one might conclude they are twice as unwell as patients B and C and 1.5 times as unwell as patient A

Rather than using the TSS, an alternative approach is to define similarity as a function of the **distance** between the patients, where we have shown distances as solid lines in Figure 3 for patients pairs A and D as well as B and C. We can measure the **rectilinear distance** between points by following only straight lines parallel to the axes: for example, the distance between A and D requires only moving two units parallel to the Anhedonia axis, resulting in a distance of 2, denoted *δ*(*A, D*). To move from B to C, we have to traverse 2 units parallel to the Mood axis and then a further 2 units parallel to the Anhedonia axis, resulting in *δ*(*B, C*) = 4. This is the *L*_1_ distance, sometimes called the “Manhattan” or “Taxi-cab” distance (Deza and Deza, 2013). Alternatively, one might choose the *L*_2_, shortest straight-line, or Euclidean distance shown as the diagonal dashed line between patients B and C which is less than the *L*_1_ distance. We choose the *L*_1_ distance because it emphasises, for example, to ‘move’ between patient B and C, there must be change in the Anhedonia *and* Mood domains in whole units on the original scale and as we show later, the *L*_1_ norm is in fact the TSS measured to the origin of the native space i.e. the point (0, 0).

To compute the *L*_1_ distance, we perform the following steps:

### Algorithm 1: *L*_1_ Distance

1. Define two clinical states as vectors **X** = (*x*_1_, *x*_2_, …, *x*_*d*_) and **Y** = (*y*_1_, *y*_2_, …, *y*_*d*_) where *d* is the number of items (dimensions)
  - for example, in our toy example with 2 items/axes (Anhedonia and Mood) we have *d* = 2 and patients A and C would be represented as **A** = (1, 3) and **C** = (3, 0).
2. Define a vector **Z** where each element *z*_*i*_ is the element-wise difference *z*_*i*_ = *x*_*i*_ − *y*_*i*_ which is the vector difference **Z** = **X** − **Y** = (*x*_1_ − *y*_1_, *x*_2_ − *y*_2_, …, *x*_*d*_ − *y*_*d*_)
  - for patients A and C we have **Z** = (1, 3) − (3, 0) = (−2, 3)
3. Define a function *F* that computes the **absolute value** *F* (**Z**) which is the element-wise maximum, max(*z*_*i*_, −*z*_*i*_) – intuitively, this simply means we take the positive value of each element *z*_*i*_
  - for patients A and C, we have *F* (**Z**) = (2, 3)
4. The *L*_1_ distance *δ*(**X, Y**) is then the **sum** of the elements of *F* (**Z**)
  - the sum of (2, 3) = 5 or *δ*(**A, C**) = 5

Equipped with this, we can tabulate the pair-wise *L*_1_ distances between each patient as:

|  | A | B | C | D |
|---|---|---|---|---|
| A | 0 | 1 | 5 | 2 |
| B | 1 | 0 | 4 | 3 |
| C | 5 | 4 | 0 | 3 |
| D | 2 | 3 | 3 | 0 |

There are two important properties shown in this table:

1. the distance between a patient and themselves (the diagonal elements) is zero, i.e. *δ*(*C, C*) = 0
2. the distance between any two patients is symmetric, i.e. *δ*(*B, C*) = *δ*(*C, B*)

To compare patients – that is, to measure their similarity in terms of measurements on the instrument/scale – we can use the difference of TSS, or their *L*_1_ distances as follows:

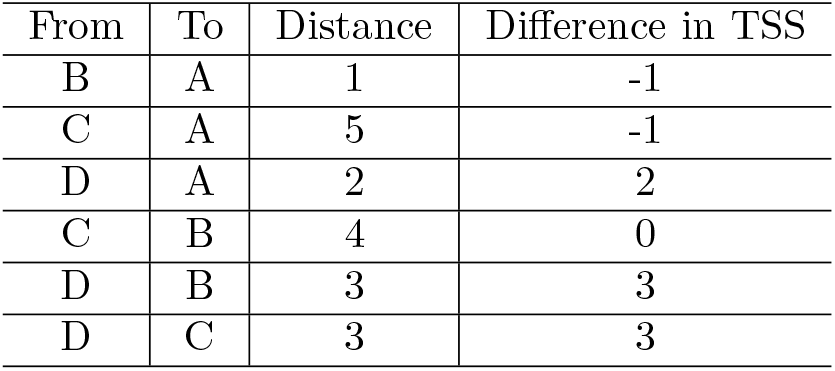

Notice that:

- Patients C and B are obviously dissimilar in Figure 3; the *L*_1_ distance respects this, but the difference in TSS is zero (this is unsurprising, because both C and B have a TSS of 3)
- Patients C and A are also obviously different in Figure 3; *L*_1_ distance respects this, but comparing them by differencing TSS gives the counter-intuitive result of -1
- Using the difference of TSS, patients D and A differ by 2; this is because the sum score for D = 6 and A = 4. However, notice that using the TSS to measure the reciprocal difference between A and D would be 4 − 6 = −2, i.e. similarity is asymmetric using TSS.

This illustrates how TSS is the arithmetic sum of *d* items that ‘compresses’ the information contained in a native multidimensional space into a convenient univariate (scalar) measure which could plausibly be used as an overall measure of severity or burden of symptoms, or to measure change in clinical state over time. The cost of this simplicity, however, is that we **lose information** about relationships (differences and similarities) between states that are clearly exposed in a geometric interpretation of the native space. Furthermore, the mapping of points from the native space to a TSS is “many to one” – many different states map to the same univariate score (cf. the subset sum problem in Section 2). Measuring distances between clinical states in the native space **preserves** information about relationships i.e. similarities and differences; using TSSs for the same purpose yields counter-intuitive results.

## 4 The Total Sum Score is the *L*_1_ Distance to an Origin

We previously demonstrated how to compute the *L*_1_ distance *δ*(**X, Y**) between two arbitrary points (clinical states): define a fixed **anchor** point **O** = (*o*_1_, *o*_2_, …, *o*_*d*_) and then measure the distance from any other clinical state (or point **X**) to that anchor *δ*(**X, O**). If the anchor is the **origin** of the native space (i.e. the bottom-left of Figure 3, indicating the minimum possible score of no symptom burden on any item/dimension), then the distance from any state to that anchor is **equivalent** to the TSS. The distance of any point measured to the origin is equivalent to the **norm** for that vector space and distance metric.

To make this concrete: set the anchor in our PHQ2 example to be the origin **O** = (0, 0) and measure the distance to this origin for each patient. The geometric result is shown in Figure 4. The *L*_1_ distances to the origin are 4, 3, 3, 6 for patients A, B, C and D respectively (i.e. identical to the TSS). This illustrates that there is a systematic relationship between vector spaces, *L*_1_ distances and the familiar TSS if we measure the *L*_1_ distance of any point to the origin.

**Figure 4:**
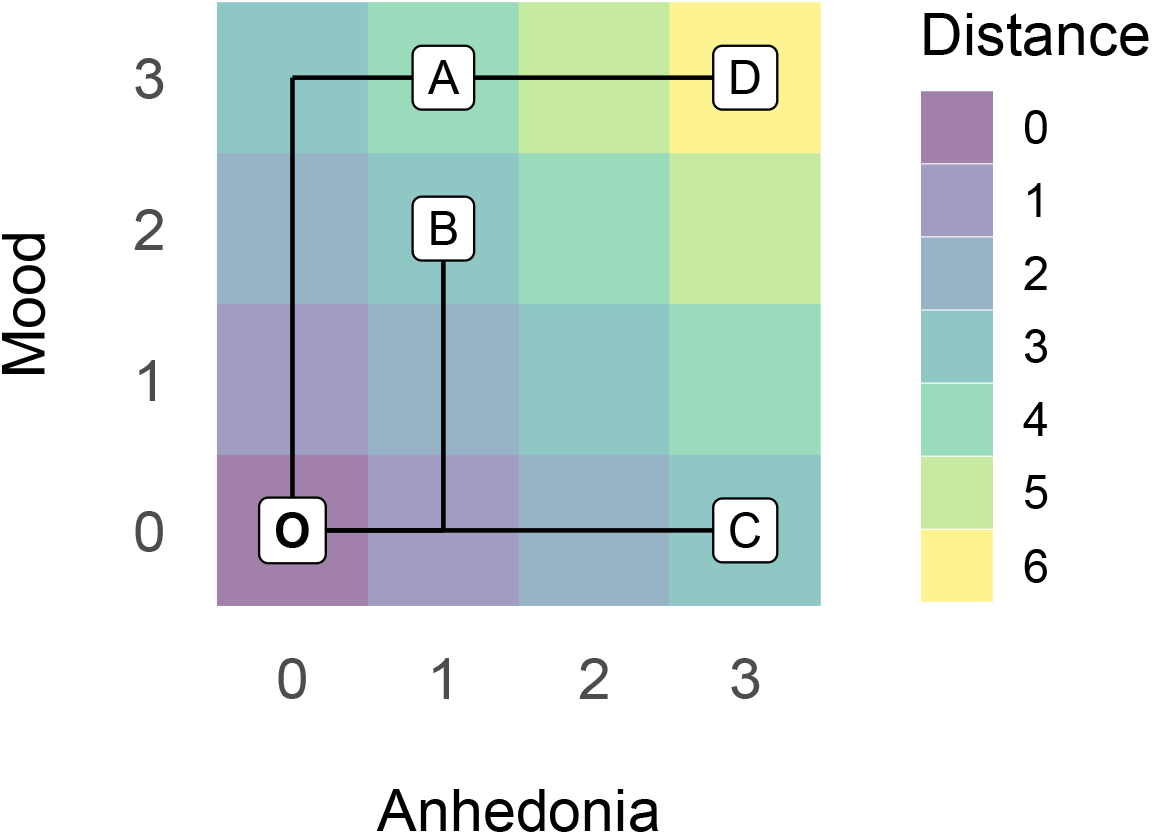
Colours represent the rectilinear distance to the origin corresponding to traversing the solid lines between patients (A,B,C and D) and the origin (O)

## 5 Anchors and Outcomes

In a cross-sectional scenario, points in the native space represent the clinical states of different patients. Alternatively, for an individual patient, each point may represent a time point in a series of measurements of clinical state. Rectilinear *L*_1_ distance measures similarities between points. In a cross-sectional setting, these distances are a measure of similarities between patients (in terms of their clinical states) and for the within-person, time series setting, the distances quantify how much a patient’s clinical state has changed between two time points.

As noted in Section 4, when the *L*_1_ distance is measured from any point to the *origin* of the native space, we obtain the familiar TSS. We have argued this is flawed because many very different points can have the same TSS. With reference to our concrete two-dimensional example:

- distance to the origin is **isotropic along each dimension/axis** – i.e. moving from a score of 3 to 0 on the Anhedonia axis has the same numerical value as moving from 3 to 0 on the Mood axis
- the rectilinear distance to the origin uses *addition* of the element-wise absolute differences (Algorithm 1, Step 4); addition is a commutative operation, such that *a* + *b* = *b* + *a*. Hence movement toward the origin (0, 0) of 2 units on the Anhedonia axis and 1 unit on the Mood axis results in a distance calculation of 2 + 1 = 3 which is the same as moving toward the origin by 1 unit on the Anhedonia axis and 2 on the Mood axis which yields 1 + 2 = 3.

If different patients (or a single patient over time) can ‘occupy’ different regions of the native space (as shown in Figure 4) then how we quantify overall severity (or change over time) could exploit geometric principles which ‘break’ the isotropy and make better use of distance metrics. In the case of the *L*_1_ distance to the origin, both improvement and severity are measured as uniform movement – along all dimensions – toward the origin resulting in the familiar TSS.

To motivate what follows, consider the case of patient-centered outcomes; the desired change or the target for treatment may be different for individual patients – for example, in Figure 4, patient C might well seek improvement in Anhedonia, but be less concerned with Mood symptoms, whereas patient A might seek the opposite. Further, rehearsing the more general problems with composite scores in the applied statistics literature (Prieto-Merino et al., 2013; Liu-Seifert et al., 2017; Troy and Simmons, 2020), we might experimentally investigate an intervention with the hypothesis that it will impact Anhedonia, but is unlikely to benefit the Mood dimension measured by the PHQ2 – so patients C and D would be expected to benefit the most, patients A and B less so or not at all.

What we propose is essentially formalising the practice of using subscales but cast in the framework of vector spaces and distances to anchors – for example, when examining negative features of psychotic disorders, our attention might reasonably be focused on items N1 through N7 of the PANSS scale and we might even discard the other 23 items corresponding to positive and general psychopathology. This subscale approach can be formalised using anchors. As a concrete example, (Andreasen et al., 2005) proposed specific subsets of the PANSS and BPRS instruments to assess response to treatment in terms of a specification for **remission**. In the framework proposed here, the subset of items becomes a specification for an anchor that is translated (moved, or shifted) away from the origin of the native space.

Returning to our two-dimensional example and using the concrete idea of remission, say we are interested in quantifying patients who have low burden of Anhedonia (i.e. 0 or 1) but where we will tolerate a higher burden of Mood symptoms (for example, Mood greater than or equal to 2). This corresponds to an anchor **O** = (1, 2). In Figure 5, the left plot shows the location of each patient on only the Anhedonia axis (similarly, the right plot shows only the Mood axis) and the red lines are the components of the anchor **O** for the Anhedonia and Mood axes respectively.

**Figure 5:**
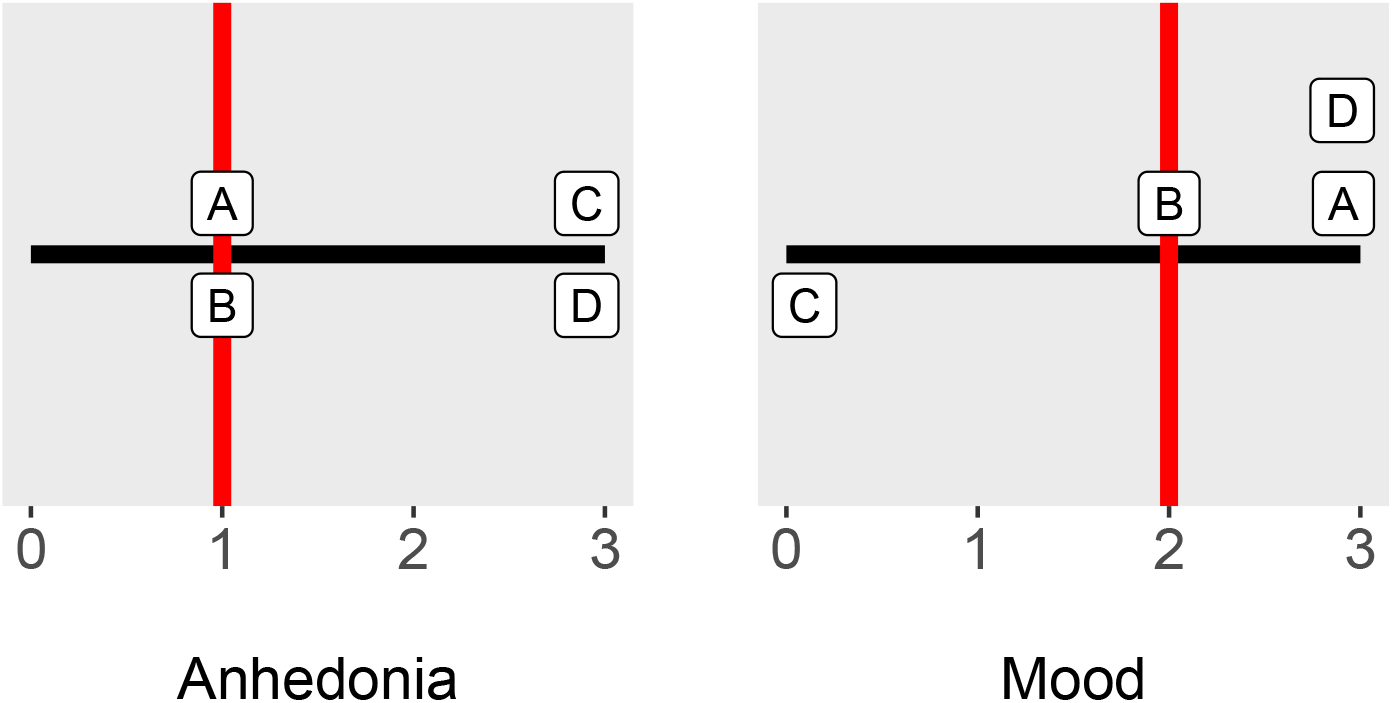
Anchors in Each Dimension

The intuition here is that:

- On the Anhedonia dimension, patients A and B are relatively unaffected, and would qualify as being in remission, but patients C and D are significantly affected
- On the Mood dimension, patients A, B and D are just above and in remission, whereas C is clearly in remission

Our proposed univariate summary should capture these properties.

We now measure the distance for each patient to the anchor (left panel in Figure 6). Notice that measuring the *L*_1_ distance from each of the same patients (A,B,C and D) to the anchor **O** changes the assignment of the composite, total score (colour) to each location. However, the distances for *each point* in the native space remain isotropic around **O**; for example, the points (1, 1), (0, 2), (2, 2) and (1, 3) all have the same distance to **O**.

**Figure 6:**
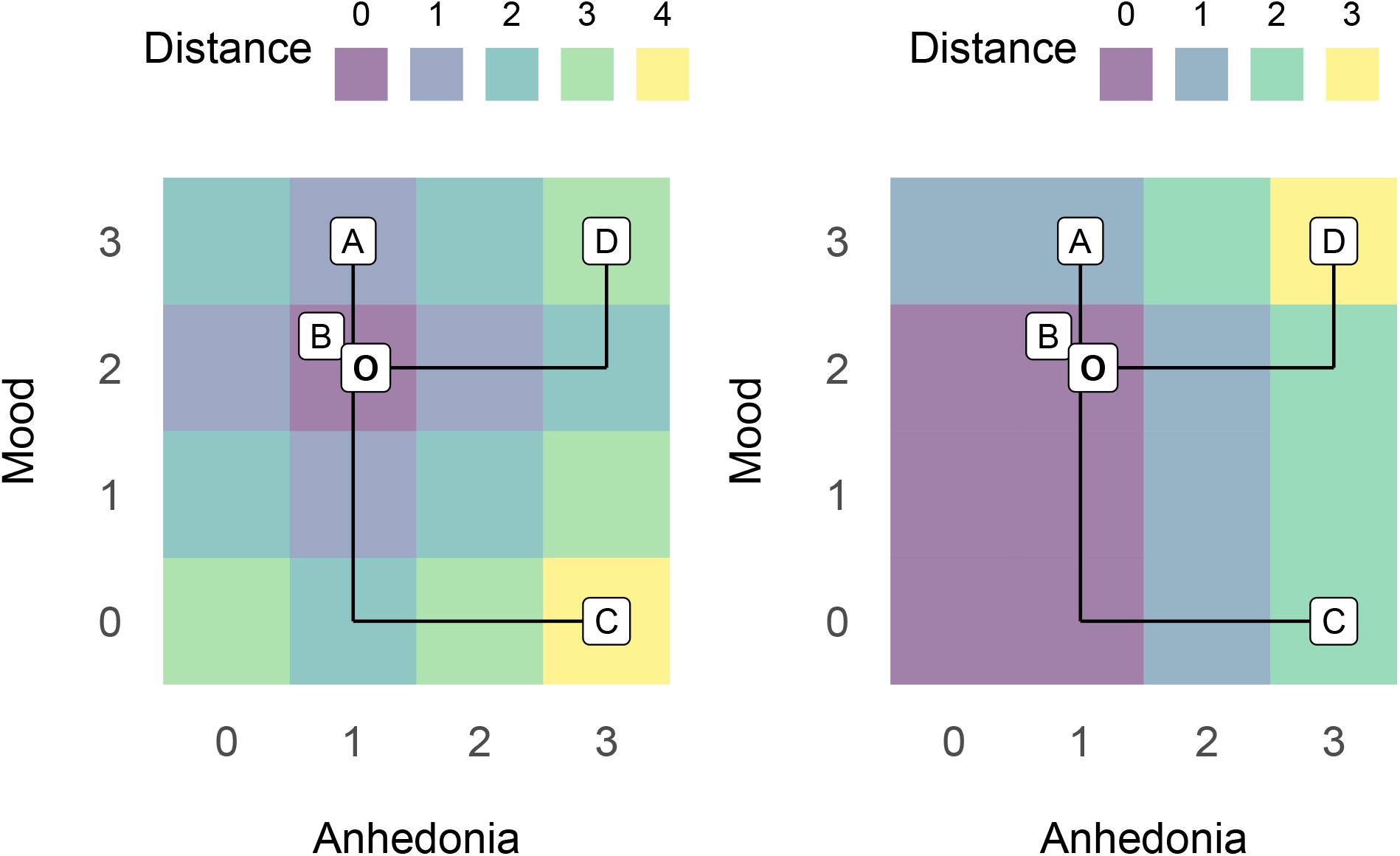
Distance to remission anchor

What we desire is that: if a patient has *both* Anhedonia ≤ 1 and Mood ≤ 2 they should obtain a score of zero indicating remission (i.e. breaking the isotropy around the anchor). We can achieve this by modifying Algorithm 1 as follows:

### Algorithm 2: Rectified *L*_1_ Distance to an Anchor

1. Define a clinical state as the vector **X** = (*x*_1_, *x*_2_, …, *x*_*d*_) with an anchor **O** = (*o*_1_, *o*_2_, …, *o*_*d*_) where *d* is the number of items (dimensions)
  - for example, in our two-dimensional example: patients A and C would be represented as **A** = (1, 3) and **C** = (3, 0). The anchor is **O** = (1, 2)
2. Define a vector **Z** where each element *z*_*i*_ is the element-wise difference *z*_*i*_ = *x*_*i*_ − *o*_*i*_ which is the vector difference **Z** = **X** − **O** = (*x*_1_ − *o*_1_, *x*_2_ − *o*_2_, …, *x*_*d*_ − *o*_*d*_)
  - for patient A, we find **Z** = (1, 3) − (1, 2) = (0, 1) whereas for patient C we have **Z** = (3, 0) − (1, 2) = (2, −2)
3. Take the **rectified value** *R*(**Z**) which is the element-wise max(*z*_*i*_, 0) – intuitively, this means any element *z*_*i*_ ≤ 0 is set to 0
  - for patient A we have *R*(**Z**) = (0, 1) whereas for patient C we have *R*(**Z**) = (2, 0)
4. The rectified *L*_1_ distance *δ*_*R*_(**X, O**) is then the **sum** of the elements of *R*(**Z**)
  - for patient A, we arrive at the sum of (0, 1) = 1 and for patient C the sum of (2, 0) = 2

This modified distance is shown on the right of Figure 6. We then find the following univariate assignments:

- Patient A scores 1 (near remission)
- Patient B scores zero (in remission)
- Patient C scores 2 and requires only a reduction of 2 units along the Anhedonia axis to be “inside” the purple region which signifies remission
- Patient D scores 3 and requires *both* a reduction of 2 units along the Anhedonia axis as well as 1 unit along the Mood axis to be “inside” the remission area

Observe that any patient inside the purple region would be assigned a score of 0, capturing the idea that differences that are considered clinically irrelevant (by definition of the anchor) can be specified. Also note that, for example, if patient C had higher Mood symptoms, it would not alter their overall score because we considered Mood ≤ 2 to be the relevant criteria for remission in terms of Mood symptoms.

The key modification to the algorithm for *L*_1_ distances to arrive at the anchored univariate score is:

1. Define an anchor that specifies clinically interesting or relevant dimensions (this essentially defines a threshold value for each dimension)
2. Modify Step 3 of the algorithm to use the **rectified** value max(*z*_*i*_, 0) instead of the **absolute** value, max(*z*_*i*_, −*d*_*i*_). We note that in implementing this modification, Algorithm 2 provides a quasi-distance metric (Deza and Deza, 2013) because the symmetry property (see Section 3) no longer holds – the implication is that one must be careful to specify that the score *δ*_*R*_(**X, O**) is the distance *from* a patient *to* the anchor and may not be equal to *δ*_*R*_(**O, X**).

Observe that the construction of the above example assumes that *lower* scores represent closeness to remission (with higher scores being further from remission) and it is trivial to define the opposite (i.e. where *higher* scores are required to move closer to the anchor – which we demonstrate later).

A final – and important – property of using an anchor and the rectified *L*_1_ distance to define a univariate score is that rather than a dichotomised “remission” criteria we instead arrive at a continuous measure of **how close** to remission any given patient is. In fact, there are many candidate functions for *R* which produce different ‘shapes’ for how the univariate score is composed (by addition) and for reasons of brevity, we have omitted a treatment of ‘weighted’ dimensions where for example, one might consider movement along a certain set of axes to carry more ‘weight’ than others.

## 6 Multidimensional Examples

We have demonstrated that for a multidimensional instrument, there are many different combinations of items that give rise to the same univariate TSS. We have then shown how the formalism of *L*_1_ distance in the native space explains why this occurs, and offer a solution in terms of defining anchors in the native space. We now support these theoretical arguments by applying them to real-world data.

### 6.1 PANSS

We extracted the pre-randomisation PANSS for each of the available 1446 patients in the CATIE trial data set (Lieberman et al., 2005) – this represents a cross-sectional sample of patients each represented in a *d* = 30 dimensional native space (i.e. where each axis represents an individual item in the PANSS instrument scored in the range 1 through 7). As is standard practice, we subtract one from each PANSS item so that the range is 0–6.

To reproduce the analysis in Section 2:

1. For each patient, we compute their TSS (i.e. over the whole PANSS instrument) which will range from a theoretical minimum of 0 (the lowest possible PANSS score) to 180 (the maximum possible PANSS
2. score corrected for each item ranging from 0–6). We find that, for the pre-randomisation data in CATIE, we have sum scores ranging from 1 to 110
3. For each TSS in the range [1, 110], we identify the subset of 1446 patients with this TSS; for example, for a TSS = 50 there are *N*_*T SS*=50_ = 39 patients
4. For the subset of *N*_*T SS*_ patients we compute
  a. the number of patients who have *identical* PANSS vectors – if TSS are perfect representations, then all *N*_*T SS*_ patients sharing that TSS should have the same PANSS vector
  b. the number of patients with *different* PANSS vectors – the higher this number (as a proportion of *N*_*T SS*_), the more patients with different vectors are being assigned the same TSS
  c. for patients with *different* PANSS vectors – we record the average number of items that are different as a proxy for the ‘diversity’ of different vectors yielding the same TSS

In the 1446 patients, there were no patients with identical PANSS vectors for *any* TSS in the sample. Figure 7 shows that:

**Figure 7:**
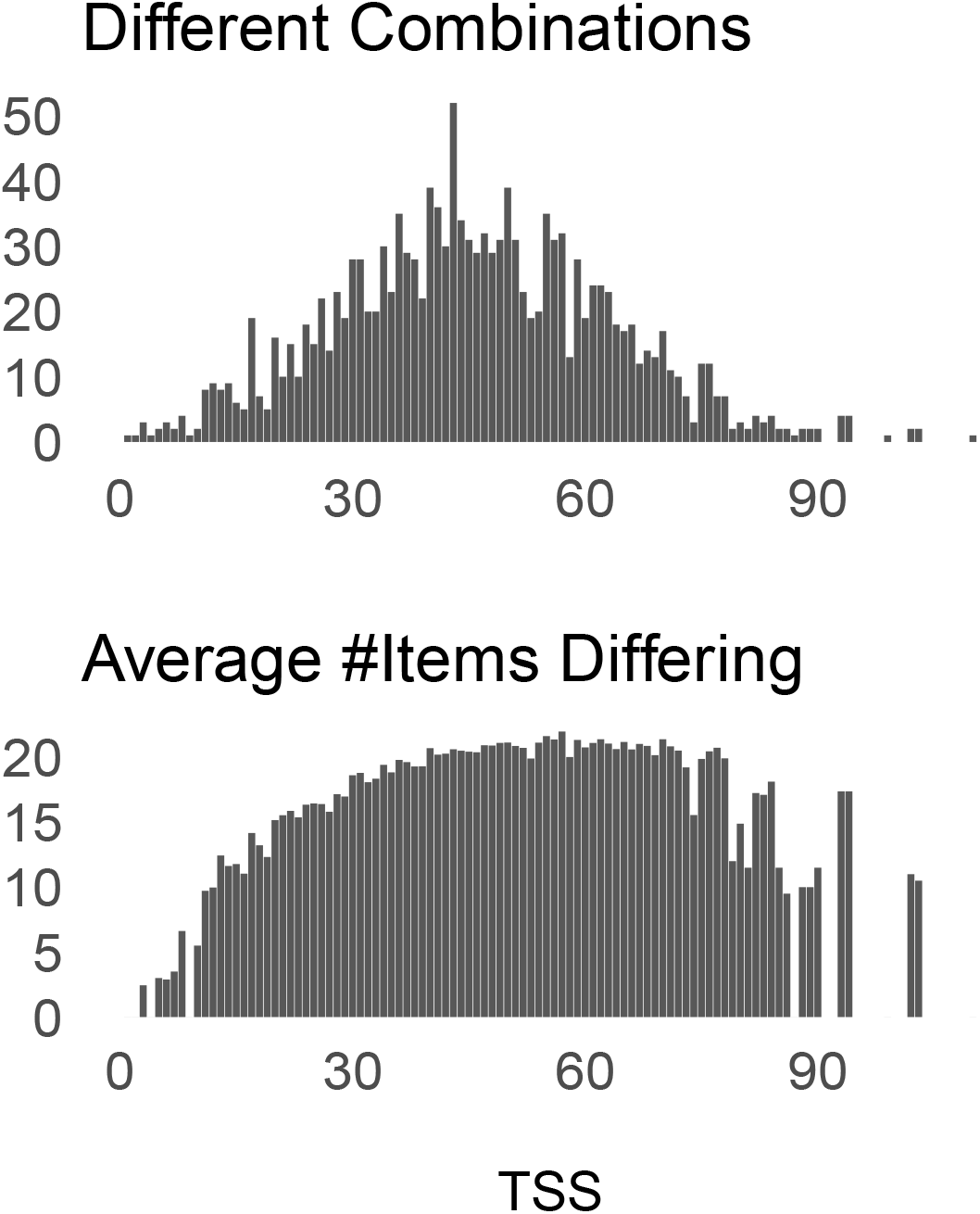
Combinations and TSS in the CATIE pre-randomisation sample

1. As for the theoretical case in Section 2, the top panel shows that the highest number of different combinations occurs around the mid-range of TSS
2. In the bottom panel, we find that, generally, for higher TSS, more items in the PANSS vectors differ; for example, when the TSS is 60, on average, those patients differed on around 20 (of a total of 30) PANSS items

### 6.2 Anchors for Remission

Having established the theoretical problems with TSSs are evident in actual data, we consider how to apply the concept of anchors to obtain alternative composite scores. An anchor should specify a desired clinical goal. If we are interested in patients’ remission status we could reasonably use the (Andreasen et al., 2005) criteria, which are: PANSS items P1, P2, P3, N1, N4, N6, G5 and G9 scoring mild or less (i.e. less than or equal to 3 on the original PANSS scale of 1–7, or 2 when each item is ‘corrected’ so that values range 0–6).

Each patient’s vector will look like:

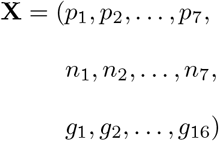

And the anchor will be:

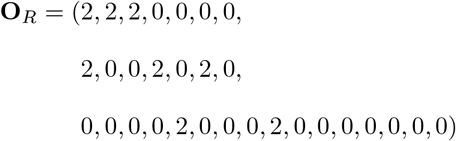

For each of the 1446 patients assessed in the CATIE pre-randomisation, Figure 8 (left panel) shows the calibration of the TSS against the equivalent distance to remission defined using the anchor **O**_*R*_ above. If the remission distance simply recapitulated the TSS, all points would be aligned on the diagonal dotted line (or similarly, all be located on a line exactly parallel to the diagonal). Instead we find a non-linear shift to overall lower remission distance scores and a ‘spread’ of remission distances for patients with the same TSS.

**Figure 8:**
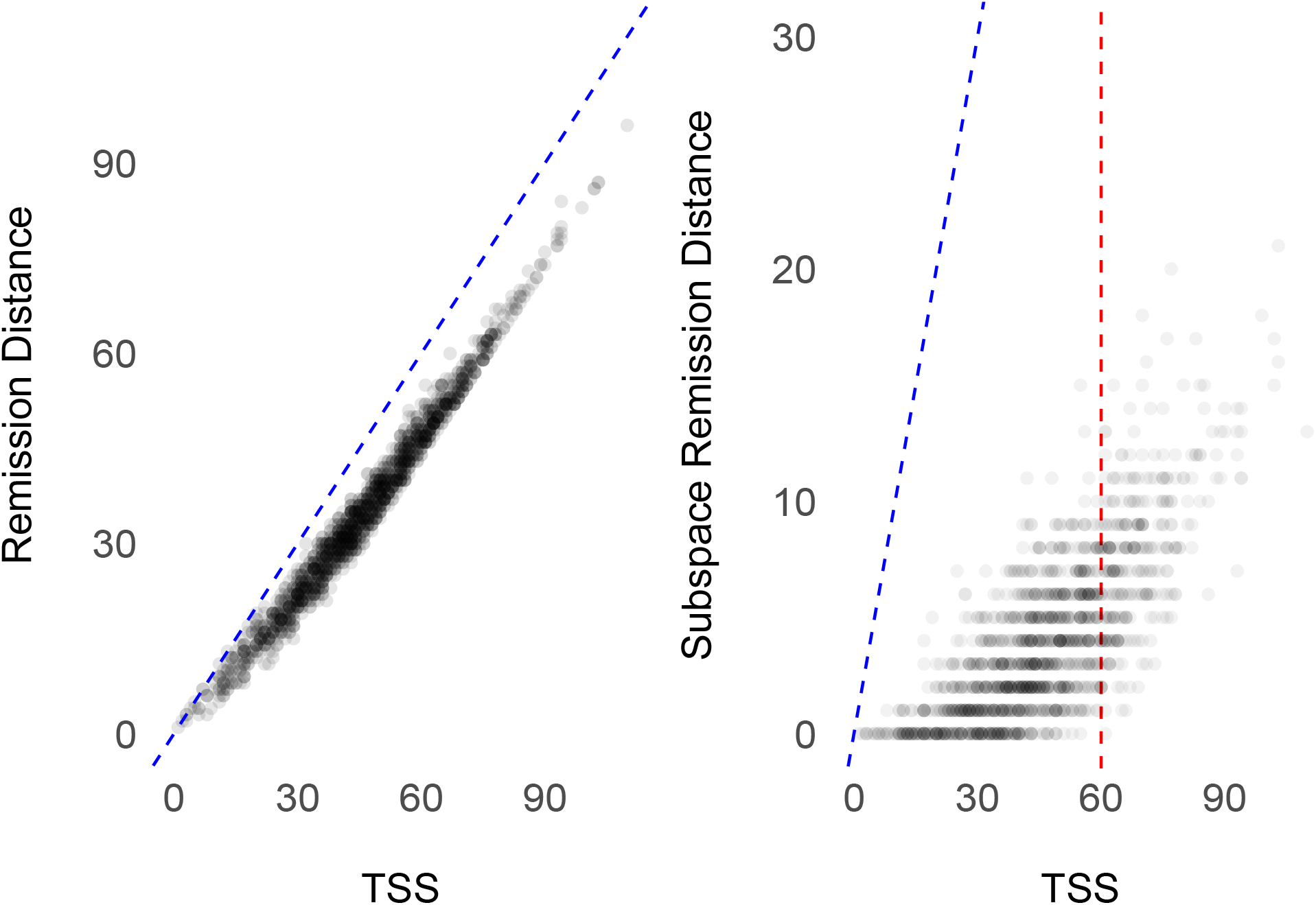
Calibration of TSS and Distance-to-Remission in the CATIE pre-randomisation sample; Blue dotted line represents the line of perfect calibration between remission distance and TSS

For each patient shown in Figure 8, their remission distance is composed of i) a contribution from the distance to the non-zero dimensions in **O**_*R*_ and ii) a contribution from the *remainder* of the PANSS items which are zero in **O**_*R*_. One could choose to isolate only the 8 anchor items, which is formally, a subspace of the *d* = 30 dimensional native space essentially discarding items in the PANSS score which are not directly relevant to the remission criteria. This is shown in Figure 8 (right panel) where patients on the “zero” horizontal line are those that (irrespective of the TSS) are in remission at pre-randomisation (by the Andreasen-criteria anchor **O**_*R*_) and patients with non-zero remission scores are represented by a continuous value representing closeness-to or distance-away from remission. The dotted red line shows that there are a broad range of differing remission scores for those patients all having the same TSS of 60.

### 6.3 Anchors for Treatment Refractoriness

We might also be interested in examining if a patient with schizophrenia has a cross-sectional clinical state profile suggestive of treatment refractory schizophrenia (TRS). One proposal (Ortiz et al., 2020) for an efficient screening for treatment-refractory schizophrenia is to measure the sum of PANSS items P2, N5 and G9 items and to then define a *positive* case (of TRS) by dichotomising at some operating threshold over this sum. Consistent with our approach, rather than a simple sum score of these items and a dichotomised ‘outcome’ (TRS or not), we will use an anchor located at the maximum value of P2, N5 and G9 and measure distances for each patient to the anchor. The required anchor is:

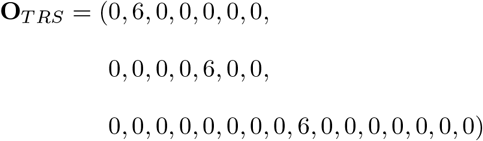

Here, moving toward the anchor **O**_*T RS*_ requires *higher* scores (in contrast to lower scores for remission). Algebraically, this simply requires that we multiply the patient vector (**X**) and the anchor (**O**_*T RS*_) by -1 when applying Algorithm 2; that is, we will measure *δ*_*R*_ (−**X**, −**O**_*T RS*_). The resulting distance will be zero when the patient strongly meets the criteria specified by the anchor, and at it’s maximum value when they absolutely do not. We will focus only on the subspace defined by items P2, N5 and G9 (that is, rather than ‘inflate’ the scores by including all 30 items).

In Figure 9 we show a 2D histogram of remission distance (i.e. the scores for patients in Figure 8, right-panel) and TRS distance for the 1446 patients in the CATIE pre-randomisation sample. The green point labelled in the top-left represents patients who are furthest from the TRS anchor (i.e. according to the Ortiz *et al* criteria) and in remission by the Andreasen criteria. Conversely, the red point in the bottom right identifies patients who are furthest from remission but closest to the TRS anchor. Intuitively, a majority of the patients who are closest to remission are most often some distance away from having a ‘profile’ identifying them as TRS – this is shown by the clustering of patients around 13–16 on the TRS axis at a remission distance of 0. If distances to TRS and remission anchors were simple transformations of each other – i.e. measured the same property – then all patients would be aligned in the histogram bins on, or parallel, to the blue diagonal line.

**Figure 9:**
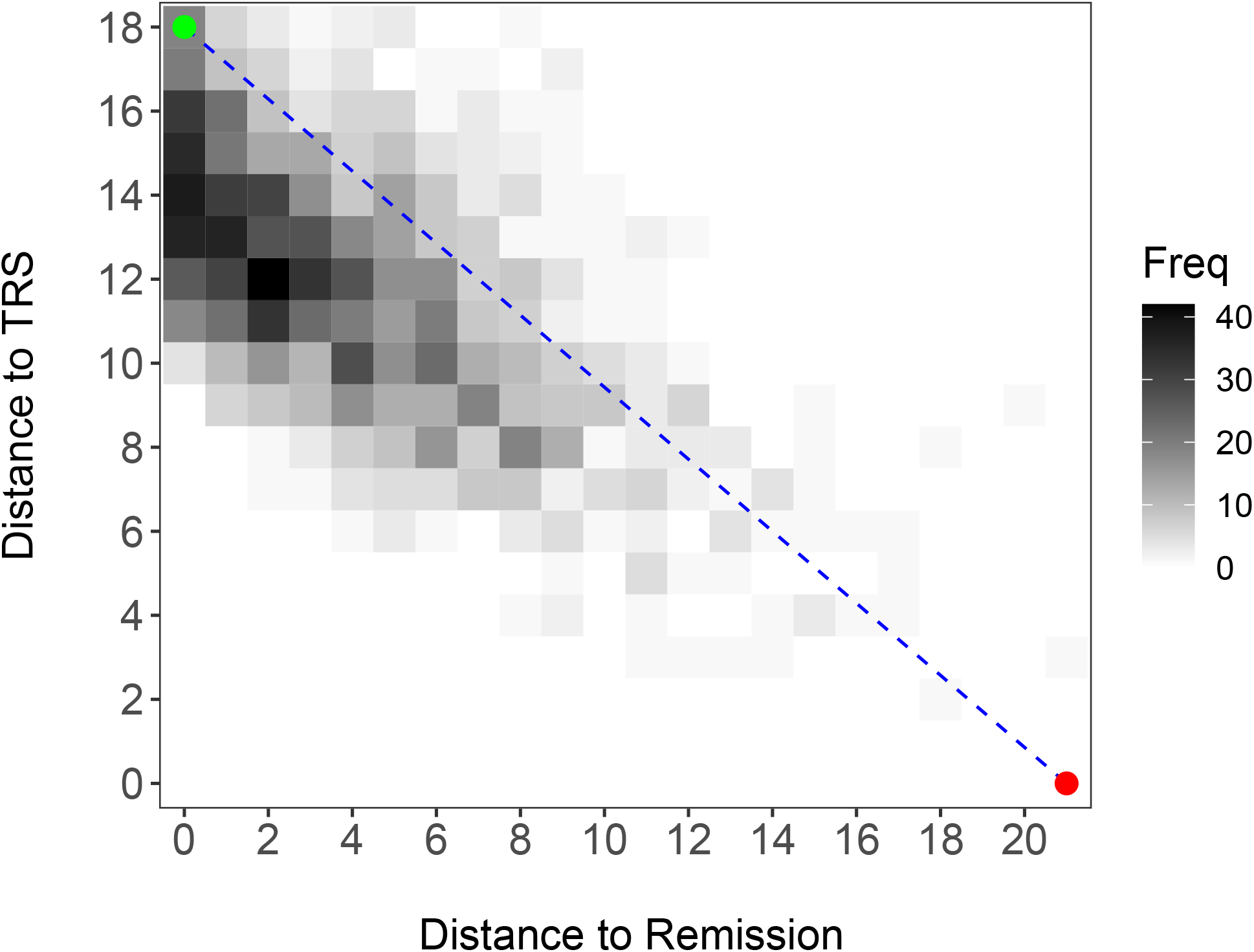
Distance to Remission and TRS Anchors for the CATIE pre-randomisation sample

## 7 Conclusion

Using informal combinatorial arguments and geometric concepts, we have argued that TSSs obscure detail in data from commonly used, multi-dimensional psychometric instruments, and fail to capture important differences between patients who clearly vary in clinical status. Related arguments against composite scores have been advanced in the applied statistics and psychometrics literature.

By conceptualising a measurement from an instrument in a *d*-dimensional normed vector space, we have illustrated how the TSS is equivalent to measuring the rectilinear or *L*_1_ distance from any location to the origin (Algorithm 1). We then generalised this by introducing **anchors** that define a clinically-relevant region in the native space and deriving a composite score by measuring rectified distances to these anchors (Algorithm 2). We propose that using distances and anchor-points retains the continuous nature of clinical phenomena, and avoids, for example, dichotomising patients into a “remission” and “non-remission” groups.

Fundamentally, this approach formalises the common procedure of using sub-scales of common clinical scales/instruments, but is not a panacea to every problem inherent in working with multidimensional instruments (Faravelli, 2004); nor does it aim to replace methods that model the dynamics of psychopathology using network methods (Robinaugh et al., 2020; Borsboom et al., 2019). Rather, we propose that when a validated instrument has utility in measuring clinical state – but we require a composite, summative score – attending to the multivariate structure of the instrument at the same time as defining what we mean by improvement, deterioration or optimal ‘states’ can be achieved with a straightforward modification to the algorithm for TSS.

In the first instance, this approach should be applied to existing datasets, both cross-sectional and longitudinal, to confirm that further clinical insights can be extracted using these methods. We propose that our approach would be particularly suited to patient-reported or personalised, patient-centric outcomes / endpoints because anchors naturally specify the important treatment targets and provide a continuous measure of how “near” or “far” a person is to it. Further work should explore a) how different functions *R*(·) in Algorithm 2 can be utilised to differentially emphasise and weight elements (axes, dimensions, items) for given psychometric instruments and b) the implications for prospectively-collected multivariate time-series data.

## Data Availability

All data used in the present study are available from the NIMH NDAR archive (with access being granted by application to the NIMH).
https://nda.nih.gov/get/access-data.html

## 8 Acknowledgements

The authors thank the National Institute of Mental Health for making the CATIE (https://nda.nih.gov/edit_collection.html?id=2081) trial data available.

